# Food for frailty: Views of older adults on development and uptake of a food-based frailty supplement

**DOI:** 10.64898/2026.04.01.26348969

**Authors:** Basharat Hussain, Ana Valdes, Stephen Timmons

**Affiliations:** School of Medicine, University of Nottingham, UK; Nottingham University Business School, UK

**Keywords:** older adults, frailty, food supplement, co-creation, healthy ageing

## Abstract

**Objective:** Frailty is an important concern in old age. Inflammation can cause frailty. Anti-inflammatory food supplements can play a role in slowing down frailty processes and consequences. This study explored the views of people (aged 50-89 years) on the need to develop a frailty supplement, preferences for its form and how older people could be encouraged to use such a supplement.

**Design:** We conducted semi-structured qualitative interviews and used a framework method to analyse the data.

**Participants:** 30 participants from a city in the UK.

**Setting:** These participants were recruited from social housing, care homes, foodbanks and the wider population. Participants were from diverse ethnic, gender and age backgrounds.

**Results:** Participants identified a strong need for the development of a food-based supplement for frailty. They expressed excitement for the supplement and viewed it as something which they would be happy to integrate in their daily food routine. In terms of preferences, our participants wanted to have multiple options, however, a biscuit-based supplement was preferred by most. The participants’ preferences were mainly based on taste of the supplement, its effectiveness, convenience in use and affordability. Muslim participants in the sample said they would be happy to use this supplement if it was developed using *Halal* ingredients.

In terms of creating awareness and encouraging people to use the proposed supplement, participants suggested a variety of marketing methods. These included: word of mouth, face to face sessions with older adults, social media, especially YouTube and advertising on TV.

**Conclusion:** The participants were generally open to the idea of a food-based supplement and felt that it could easily fit with their existing food practices and lifestyles.

**Strengths and limitations of this study:**

- This is the first study to explore views of older people for the need to develop a frailty supplement, preferences for its form and how older people could be encouraged to use such a supplement.
- Qualitative interviews allowed to collect rich data which is helpful in understanding if older people want a food-based supplement and what their preferences would be.
- Data was collected from a diverse sample in terms of settings (social housing, food banks, care homes), ethnicity, age and gender.
- Findings are based on a small sample size of older people from a single city in the UK and may not be generalizable to other areas.

## Introduction

There is no consensus on a definition of frailty (1). However it is usually understood that frailty is a state of vulnerability where a human body is less resilient in coping with a stressor event (2). Frailty has many negative consequences for health and is associated with poorer health outcomes and quality of life among older adults. Compared to a non-frail group, frail older adults were more likely to be hospitalised and have a significantly increased risk of dying within 3 years (3). Moreover, frailty also results in a 1.6 to 2.0 fold risk for loss of daily living activities; and a 1.2 to 2.8 fold risk for falls and fractures (4). These statistics suggests that frailty is an important public health concern.

A systematic review and meta-analysis of population-level studies on prevalence of frailty in 62 countries across the world found that the prevalence of frailty is 13% to 24% (5). The review also noted that frailty is unequally distributed; with prevalence of frailty higher among females (15% to 29%) than males (11% to 20%)(5). Another systematic review and meta-analysis found a higher prevalence of frailty in nursing homes compared to older adults living in community (6). It is also noted that older adults from a poor socio-economic background are more likely to become frail (7). These statistics are likely to increase with ageing populations worldwide.

Research has shown that inflammation in the body play an important role in causing frailty (8, 9). Malnutrition is a major risk factor for inflammation and the development of physical frailty among older adults (10). Nutritional supplements containing fibre, calcium, protein and vitamins are found to be helpful in preventing and/or reversing physical frailty among older adults (11). However, ensuring that older adults accept these nutritional supplements and integrate these into their daily food routine is a big challenge (12).

A qualitative meta synthesis showed that nutritional interventions that meet individual preferences and that can easily be integrated with the preexisting lifestyles of older adults increase their uptake (6). Older adults prefer supplements which have proven health benefits, contain natural ingredients (13) and are affordable(14). Food-based supplements such as protein-rich yogurt can easily be incorporated in the diet of older adults (15). Additionally, the sensory properties of the supplement (e.g. taste, texture and appearance) are also important for older adults(16). For example, in a sip-feed nutritional supplement study among older adults(17), the participants preferred vanilla and apple, whereas chocolate was least preferred. As the perceived sweetness of an oral nutritional supplement increases, preference for the supplement decreases among older adults (18). Savoury flavoured supplements such as coffee and tomato are preferred by older adults (19).

The format of a nutritional supplement is also an important factor that contributes to its uptake. Developing supplements that can be readily incorporated into the everyday lives of older adults enhances their uptake (20). Soup is an important part of daily food routines among Portuguese and Spanish people, therefore when older adults living in care homes in these countries were offered a protein-rich kale soup which was well accepted (21). A review has found that a liquid format for nutritional supplements can enhance their acceptability among older adults (22). Food scientists are also looking into potential of developing a functional biscuit that can be offered as a protein supplement (23).

People make food choices to express their identity and cultural values (24); therefore, variations in cultural preferences were identified as having the most influence on preferences for fortified meals among older adults in different European countries (25). Similarly, the religious identity of consumers also plays a role while making decisions about what to eat and drink (26). For example, Muslim consumers would consume a nutritional supplement if it was made of Halal ingredients (27). Hence, it is important that when a nutritional supplement is developed, the process and ingredients used need a transparent explanation in order to improve uptake (28).

The packaging of a food product is also an important factor in determining its acceptability among older adults. An study conducted in Australia explored hand and finger strength of older adults in opening various food packs (29). Factors studied included: the attempts made at pack opening, the time taken to open the pack and its correlation between grip and pinch strengths with opening. The results from the study found that the most difficult packages to open included: tetra packs, water bottles, cereal, desserts, fruit cups, biscuits and cheese portions. Therefore, nutritional supplements that are in easy-to-open packaging are more likely to be accepted by older adults (30).

It appears that there are variety of preferences among older adults in terms of taste, format and packaging of a food supplement. Therefore, a more person-centred approach which considers individual needs and preferences is needed for development of a supplement for frailty (31). Offering a variety of flavours and formats (e.g. soups, sweet drinks, biscuits) could be helpful to improve appeal and therefore uptake (31). Meeting individual preferences and needs is challenging for companies developing nutritional supplements (13). It also raises the question whether poorer older adults would be able to pay the cost of supplements (32). Therefore, support from publicly-funded schemes (such as the UK NHS) would make supplements more accessible (33).

Researchers can involve older adults in the development, promotion and evaluation of new nutritional supplements for frailty. Co-creation of nutritional supplements has been well received across a range of nutrients, including fibre and protein (34). Through involving older adults in the process, the researchers benefited from the lived experiences of older adults (35) and had a better understanding of the preferences of older adults for the development, promotion and integration of these supplements into daily routine of the older adults. However, involving older adults in the development of these supplements is challenging (36). Challenges include the recruitment of older adults from a variety of settings, their time, and a lack of structure for the co-creation process (36).

Healthcare professionals can intervene to improve the acceptability of nutritional supplements in older adults. Research has found that given that older adults place considerable confidence in their medical advice (37). Therefore, trusted nutritional advice, for instance from a recognised healthcare professional (e.g. GPs, dietician), also increases the acceptability of nutritional supplements among older adults(15). Healthcare professionals can play their role by endorsing the product to the older adults and conveying its positive benefits for health(38) . However, for healthcare professionals to endorse a nutritional supplement, the supplement should be evidenced-based, and it should deliver what it claims on the label (30). Older adults living in institutional settings such as care homes have to eat what is on offer (39), therefore, staff in care homes can play an important role through offering a range of food options and assisting older adults to take these.

Along with researchers and healthcare professionals, older adults themselves can also play a role in the uptake of a frailty supplement. Raising older adults’ awareness of the need for a nutrient such as protein in ageing can also contribute to its uptake (25). During awareness raising efforts, food-based examples should be given in a clear and accessible format (e.g. a factsheet as well as practical tips)(40). Older adults are not a homogeneous group in terms of their ethnicity, daily food routines, therefore segmenting them into groups and then implementing relevant educational and public health strategies would help increasing their awareness about the need of and acceptability of a nutritional supplement (41).

This study aims to explore views of older adults for the need and preferences of a food-based supplement for frailty.

## Method

We used a qualitative approach to undertake this study. Ethical clearance for the study was obtained from the Nottingham University Business School Research Ethics Committee. All participants gave informed consent.

A purposive sampling approach was used to recruit the participants. We recruited 30 participants from social housing, care homes, foodbank users and the wider population. Based on the research questions and objectives, a topic guide was developed. This topic guide included questions around existing daily food routines, factors affecting these routines, and whether participants were taking any specific food or supplement for health. Participants were also asked if they thought there was any need for a food supplement which could help them to live well; and how this food supplement should be formulated and made available to encourage uptake among older adults. Using the topic guide, BH conducted face to face and online semi-structured interviews. BH is an experienced qualitative researcher and has researched food practices previously. All interviews were digitally recorded.

Recorded interviews were transcribed verbatim by a professional transcription service. The framework method was used for data analysis (42).

## Findings

We interviewed 30 participants: 19 from the general population, 04 from social housing, 4 from care homes and 3 from users of a local food bank. 16 participants were male and 14 female. In terms of ethnicity, 10 were British Pakistanis, 7 White British, 6 Black African, 4 British Bangladeshi, 2 British Indian, and 1 mixed race.

The age profile of the participants was: 50-60 years (n:17), 61-70 years (n: 7), 71-79 years (n: 3) and 80-89 years (n: 3)

### Theme 1 Existing food routine and factors affecting it

Participants usually ate three times a day; breakfast, lunch and dinner. Snacking was also a part of daily food routines. There are number of factors affecting food routines.

For example, some participants reported that as they were getting older, they were more cautious about their food choices.

> *I think it’s important to stay health conscious, to have a variety of food. … especially as you get older. When I was younger, I didn’t care what I ate. (Age 60s, Female, British Pakistani, General population)*

Food taste was an important factor.

> *I think I eat food which I enjoy eating. I need to enjoy the food first and health is next thing. (Age 50s, Male, Black African, General population)*

Living arrangements were also found to be linked to food choices. For example, living in a care home affected choices strongly:

> *It’s very hard eating away from your own home. You have what you want at home. Here [in the care home] they would say the choice is jacket potato or an omelette. Because they have to cater for large population. (Age 80s, Female, White British, Care home resident)*

Participants also reported that their health conditions affected food choices.

> *I’ve just recently been diagnosed with coeliac disease so that’s changed my diet a lot. Now I can’t have chapattis, I can’t have bread, I can’t have cereal either, it has to be gluten-free cereal. (Age 50s, Female, British Pakistani, General population)*

Most participants reported that their financial situation limited their food choices. Participant 29, from the foodbank users group, reported:

> *Normally, like, just simple things like breads, pastas, rice because it’s cheaper food to buy. [For lunch] If I don’t have enough money, I have toast and butter. … Maybe ham and cucumber because … you can get ham cheap £1*.*50 [Dinner] Maybe like a ready meal, like a pasta ready meal for example, just-I don’t know, it just depends on what’s cheap. (Age 50s, Male, Black African, Foodbank).*

During the interviews we asked participants if they were taking any specific food or supplement for health reasons as part of their daily routine. Those who were taking supplements were usually advised by their doctors.

> *I’ve just started taking iron tablets but that’s a prescription from the doctors and that’s when I got diagnosed with coeliac disease. (Age 50s, Female, British Pakistani, General population)*

### Theme 2 Views on idea of development of a food supplement for frailty and intention to uptake

Participants across genders, ages, ethnicities and settings (e.g. care homes, foodbanks, social housing) noted that currently there is an unmet need for a food-based supplement which is effective in preventing and reducing ageing-related issues such as frailty, or aches and pains. Participants were unhappy with their pain medication and were looking for something else. A participant from the social housing group noted that ‘*falls can happen to anybody at any time as you’re getting older, I want to maintain my independence as much as possible. Anything really that dietary wise would be good’* (Age 50s, female, White British).

When asked about their intention to use a proposed supplement, all the participants responded ‘Yes’. However, some participants also said that their decision to use this supplement will be conditional on being advised by a healthcare professional, its price, being effective, evidence-based, having no side effects and using acceptable ingredients.

> *If the doctor advised me then I can take it, otherwise I wouldn’t eat it. (Age 60s, Female, British Pakistani, General population)*

Participants from the Pakistani and Bangladeshi population groups highlighted that they would take this supplement if the ingredients used were Halal.

> *The Muslims are very sensitive about halal and haram especially living in the UK. So, these are things that are very important. (Age 50s, Male, British Pakistani, General population)*

### Theme 3 Preferences on form of frailty food supplement

There were some participants who said they *‘don’t mind’* about form of the supplement as long it was effective or prescribed by their doctors.

> *It’s not a problem with a biscuit and powder and liquid, if that supplement is good, I don’t have any problem. (Age 70s, Male, British Bangladeshi, General population)*

Participants preferring a biscuit gave several reasons. Compared to a powder, a biscuit is something that most people are already used to and is convenient. They thought that a biscuit-based, instead of a tablet-based-supplement would be psychologically more acceptable to people as they would think that they are taking a food, rather medication.

> *Because I eat lots of biscuits with tea, anyway, and they will make it tasty as well. (Age 50s, Female, British Pakistani, Social housing group sample)*

> *Biscuit would be a very good idea, because I’ve noticed a lot of older people, they don’t eat meals, they will have tea and biscuits. So, if you can implement something in biscuits for them they’re gonna have the biscuits be comfortable but they’re still gonna get what they need in their body, that’s a really good idea. (Participant 29, Age 50s, Male, Black African, Foodbank).*

Some preferred cake over biscuits as it is softer than biscuits. Participants who did not preferred the biscuit-based supplement had several reasons. These included that eating biscuits would *‘kill your appetite’* and that biscuits are sweet, and this would not be helpful for people who are diabetic.

> *Biscuits are too sweet and I’m diabetic. I do not want a biscuit for my supplement. (Age 80s, Male, White British, Care home resident)*

The second most preferred option among the participants was a liquid-based food supplement. Participants’ rationale for this option was that the liquid form of supplement would be *‘already prepared, ‘easy and ready to drink’*. They mentioned examples of drinks already available such as ‘Fortisip’ and ‘Paediasure’.

> *If it’s readily prepared like orange juice, you know those children orange juice that is already in the pack. That’s good. I prefer that one. (Age 50s, Female, Black African, General population).*

Although the majority of participants disliked tablets, a few participants said they would prefer tablet or capsule-based supplements. They said tablet and capsules are easy to take, especially for older adults who can forget their medication.

> *Because they’re easy to take. (Age 50s, Male, Black African, General population)*

> *I’ve got my pill box; there’s Sunday to Saturday and it’s just one compartment for each day. (Age 70s, Female, White British, Social housing)*

Participants who were not in favour of a tablet-based supplement noted that there is stigma attached to taking tablets. Furthermore, many older people are already taking several tablets so would not like to add more. There were only two participants who wanted to have the frailty supplement in powder format.

### Theme 4 Taste preferences for the food supplement

The participants reported as long as the supplement was effective and prescribed by their doctors, taste *‘doesn’t matter’* for them. Some suggested that the supplement shouldn’t be ‘*too unpleasant’*.

> *I would take anything if I have a problem of frailty … But if they make it tasty, it will be easier for anyone to consume. (Age 50s, Female, British Pakistani, Social housing)*

Some participants also wanted to make it tasteless ‘*because not everybody’s going to like the taste of strawberries or bananas or coffee (Age 70, Female, White British, Social housing group*). However, some suggested that it needed to have a good taste, as this would help in increasing uptake.

> *The majority of the stuff that we eat in our daily lives, it depends on the taste. If it tastes nice then you like to have it. If it doesn’t taste nice, you’d rather avoid it. (Participant 16, Age 50s, Male, British Pakistani, General population)*

One participant from the foodbank sample wanted salty biscuits as ‘*our bodies need a certain level of salt to be able to function’*. However, some also noted problems with a salty taste:

> *I don’t like salty stuff. I don’t even like crisps that are salt and vinegar. That’s what why I’m saying, bland. If it’s salty then you’ve got the blood pressure people who wouldn’t take it. (Age 50s, Female, British Pakistani, General population)*

### Theme 5 Views on availability of the supplement, its price and packaging

Most participants said that the supplement should be easily available in multiple places like pharmacies, supermarkets and ethnic shops.

> *I think this kind of stuff should be available in the supermarkets because to go to a pharmacy, psychologically you’re thinking you’re getting a medicine. (Age 50s, Male, British Pakistani, General population group)*

However, there were some who suggested that the supplement should be prescribed *‘through [the] GP if that is going to be the criteria to make it free’*. People in care home also suggested that it should come via the doctor.

> *My position at the moment it would be the doctor because again people in my position (in care home) can’t get out to go to the chemist, but the staff can here repeat prescription things. (Age 80s, Female, White British, Care home resident)*

Many disagreed on this option suggesting that it would add an extra step of making a GP appointment.

> *To get the GP appointment is a headache. It might be hard to get an appointment with a GP. (Age 50s, Female, British Pakistani, Social housing).*

It was noted that whatever the form is, its packaging should be easy for older people to use.

> *if you’re aiming it at elderly people it should be fairly easy, it’s just a case of ripping the top of it and you can pour it out or drink out of it. The more complicated it gets the less interesting it becomes. (Age 80s, Female, White British, Care home resident)*

Participants suggested that in case of a liquid or powder form of supplement, it should be packed in single serving so that older people did not need to bother with storage. A participant suggested whatever the packing is, it needed to be aesthetically appealing, and inclusive:

Participants found it hard to give any price, though there were few participants who wanted it to be a minimal price or even free.

> *It should be something that is very, that’s going to be very, very affordable, maybe £1 per week and even free for over 65 (Age 50s, Female, Black African, General population).*

However, most participants were happy to pay £5 per week of supply.

> *I will say maybe like £5 per week, so that it can be good for everyone (P13)*

### Theme 6 Creating awareness among older adults for the frailty food supplement

Participants suggested various ways to create awareness about the supplement to increase its uptake among the older adults. They suggested using traditional media.

> *I think television would be the best, because we all watch television. Advertise on main channels like BBC and ITV. Not newspapers as older people don’t read. (Age 80s, Male, White British, Care home resident)*

Participants suggested using videos and leaflets. Participants from ethnic minority background suggested creating awareness about the supplement in ethnic languages to help reach to the older people directly.

> *You make small videos of different people from different walks of life, which are advocating this supplement. (age 70s, Male, British Pakistani, General population).*

However, it was noted that leaflets may not be a good way to raise awareness especially among ethnic minority older adults who possibly could not read English. It was suggested that for this population, face to face events at a community level would be helpful.

> *You put a leaflet, you’re not able to read English. I think the oral communication, face to face in the community centre. (Male, 60s, British Pakistani, General population).*

To increase its uptake among Muslims, participants suggested that the supplement should be developed with halal ingredients.

> *Make sure you have Halal logo on packing, because Muslims, especially old people, they are very sensitive about halal and haram food. (Age 60s, Male, British Pakistani, General population group).*

All participants suggested multiple places where the supplement could be marketed. These include residential homes, place of worship, day centres, community centres, GP surgeries, pharmacies, libraries, cafes and foodbanks. Participants identified that healthcare professionals are considered trustworthy and older people meet them frequently.

> *They have advice from the nursing staff. They usually have carers come to their house; that’s the best people. (Age 60s, Female, British Pakistani, General population)*

Participants, especially among ethnic minority groups, suggested using word of mouth to spread awareness.

> *I think the only thing is word of mouth, (Age 50s, Male, Black African, General population)*

Offering tasting sessions would help to increase uptake.

> *If you can offer the trial to some people, and if those people would be getting real benefit, then it would be very quickly boost up (Age 50s, Male, British Pakistani, General population)*

In terms of framing of message for marketing, participants suggested including benefits of the supplement in terms of preventing frailty-related issues and curing existing issues such as pain.

> *Maybe market it as a food with medicinal capabilities. A bit like sea moss, it’s got medicinal good for your body properties, but it’s food. (Age 50s, Male, Black African, Foodbank sample group)*

In order to avoid stigma, some participants suggested avoiding using age-specific wording in the marketing messages.

> *Don’t write that it’s for older and frail people because then it is like you are stigmatising people on the basis of their age. (Age 60s, Male, British Pakistani, General group)*

## Discussion

We interviewed because food and eating are known to be strongly cultural and contextual practices, and it was important to understand our participants’ preferences in these contexts. What is encouraging about this data is that our interviewees were generally open to the idea of a food-based supplement and saw that it could fit with their existing food practices and lifestyles. This is the despite the fact that even with a relatively small (albeit diverse) sample, there is a great deal of variation in the kinds of food people eat and how they eat them (time and place). Most of our sample were aware of links between food and health.

Unsurprisingly, convenience was a significant issue (29), though what was thought convenient was different for different people. The data on cost was also encouraging – it seems like a majority of participants would be prepared to pay at least something (33). Data also illuminate possible marketing strategies and platforms, though we would favour co-design of the supplement which would make any marketing inherently much more likely to be successful (36).

Our data suggest that a co-design/co-production approach to the design, packing ad marketing approach to a novel food-based supplement is both feasible and desirable (36).

## Data Availability

All data produced in the present study are available upon reasonable request to the authors

## Acknowledgement

Thanks to all participants for their help with the data collection.

